# Acceptability of advanced Controlled Human Infection Models; the case of a pneumococcal challenge in people living with HIV and mycobacterial challenge with Bacillus Calmette Guerin in Malawi

**DOI:** 10.64898/2026.03.19.26348855

**Authors:** Anthony E. Chirwa, Charity Gunda, Shalom Songolo, Edna Nsomba, Neema Toto, Lumbani Makhaza, Mphatso Mailboy, Clara Ngoliwa, Linda Chamtunga, Modesta Reuben, Chikondi Chakwiya, Pemphero Liwonde, Andrew Sigoloti, Evaristar Kudowa, Godwin Tembo, Ben Morton, Bridgette Galafa, Faith Thole, Lorensio Chimgoneko, Brenald Dzonzi, Vitumbiko Nkhoma, Edward Mangani, Mphatso Mayuni, Alfred Muyaya, Fanny Kapakasa, Morrison P Kamanga, Tiyamika Nthandira, Diana Mwaipaya, Nengie Chirwa, Ndaziona P.K. Banda, Marc Y R Henrion, Tarsizio Chikaonda, Kondwani C. Jambo, Blessings Kapumba, Stephen B Gordon, Anna C T Gordon, Deborah Nyirenda, the Malawi Accelerated Research in Vaccines, Experimental and Laboratory Systems (MARVELS) consortium

## Abstract

**Background:** Controlled Human Infection Models (CHIM) in which volunteers are experimentally exposed to pathogens are used to study pathogenesis and down select among treatment and vaccine candidates. Scientifically, CHIM studies are best conducted among at-risk populations in which the infection is endemic, and who are likely to benefit from new interventions tested. Ethically, the perspectives of these communities should be carefully considered in designing such research.

CHIM studies are often conducted among healthy adults, but we wished to understand study participant’s and community views on conducting CHIM studies in groups other than healthy adults, particularly people living with HIV (PLHIV). We also wished to explore community perceptions of a tuberculosis CHIM (TB CHIM), initially using Bacillus Calmette Guerin (BCG) as a safe intermediate step.

**Method:** We conducted fourteen focus group discussions (FGD) and eight in-depth interviews (IDI) among a wide range of stakeholders including participants, health workers, community advisory groups, religious leaders and medical opinion leaders in Malawi. Discussions and interviews were recorded, transcribed and analysed using thematic and framework analysis .

**Results:** Four themes emerged showing a general acceptance of CHIM studies, and confidence in the research practice and motivation. In the specific examples discussed in this work – at-risk populations specifically PLHIV and complex pathogens specifically TB-CHIM, there was concern for safety, proper consent and community consultation. In addition, cultural views on gender roles in Malawi, tissue sampling and residential stays for research study were discussed.

**Conclusion:** Overall, there was support for CHIM among PLHIV, and CHIM working through a BCG CHIM towards safe TB CHIM in future. Appropriate safety caveats and regulatory measures were suggested. Advice on wide, public awareness campaign methods was offered to advise researchers.

## Introduction

Controlled Human Infection Models (CHIM, - also called human infection studies, HIS); involve deliberately introducing an infectious organism in humans to understand disease processes and immune responses and accelerate intervention development[1]. CHIMs have been utilised as a tool to answer questions on disease acquisition and transmission dynamics and have contributed to vaccine development[2, 3]. CHIMs are now widely accepted and have been conducted in both high-income, low disease-burdened settings such as the UK[4], and in low-income high disease burden settings such as Malawi[5]. Additionally, ethical and practical considerations for CHIMs have been developed and standardised[6–8]. As confidence in this method of infectious disease research grows, improvement of the models by testing at-risk populations will increase, and the range of pathogens used will expand.

Human infection studies have progressed to include at-risk populations such as a pneumococcal CHIM in elderly participants in Liverpool, UK[9], and malaria CHIM in Kenyan adults[10]. CHIMs in development include people living with HIV (PLHIV)[11]. HIV infected individuals are at increased risk of infections including pneumococcal disease and exhibit higher residual pneumococcal colonisation than HIV negative individuals[12, 13], but PLHIV remain excluded from pneumococcal vaccination programs in sub-Saharan Africa. There is a case for advancing vaccine discovery in PLHIV using CHIM, but the community acceptability of this case has not been explored.

CHIMs targeting vaccine-preventable diseases were developed initially studying bacterial infections with clear rescue strategies[2]. Viral infections without clear treatment protocols have since been studied, including influenza, dengue and SARS-CoV-2[14]. Among infectious diseases considered for CHIM, tuberculosis stands out as a particularly complex and high-risk candidate. Tuberculosis (TB) has risk and stigma, and persistent infection can be reactivated by immunosuppression in later life. CHIM using attenuated strains such as Bacillus Camille Guerin- BCG have been conducted in both TB endemic and non-endemic settings. These studies have provided useful information towards accelerating TB vaccine development[15–17]. Mycobacterial models, colloquially described as “TB CHIMs” in this work, have largely been understudied in endemic settings, where populations are most affected and the need for better vaccines beyond the licensed BCG is critical. BCG is the only current mycobacterial CHIM, but the possibility of genetically modified BCG or *M.tuberculosis* being used in future makes “TB CHIM” an appropriate topic for immediate and serious discussion[18]. Importantly, however, community views of such a model in an endemic area have not been explored. The PLHIV community are at high risk of tuberculosis and so are stakeholders in this discussion, allowing the two topics in this study to be brought together.

In Malawi, the Malawi Accelerated Research in Vaccines by Experimental and Laboratory Systems (MARVELS) consortium has been conducting CHIM research since 2017. We have established a stakeholder engagement strategy to assess the acceptability of advanced CHIMs, called ACHIMA (Advanced CHIM Acceptability). The ACHIMA programme, through public and participant involvement and engagement (PPIE) has proposed ethical and practical considerations for a CHIM in people living with HIV, and for a tuberculosis CHIM using BCG[19]. Using a consultative community engagement strategy and experience from an established pneumococcal CHIM methodology, we now present stakeholder views on both the conduct and ethics of the first controlled human infection model in PLHIV and a tuberculosis challenge using BCG in HIV negative adult Malawians.

## Methods

We conducted focus group discussions (FGDs) and in-depth interviews (IDIs) including a wide range of stakeholders to understand community views and to take advice on practical and ethical considerations for CHIMs in an at-risk population of special interest in Malawi - people living with HIV (PLHIV). We also engaged these participants to understand community views and to take advice on practical and ethical considerations for CHIM methods being applied to a disease of specific interest in Malawi – tuberculosis. Malawi is a country with 11% of urban adults living with HIV, in which tuberculosis remains a high-burden disease and a national priority.

### Participants and recruitment discussions

Potential FGD and IDI participants contacted through advertisement, public engagement structures and professional contacts were offered an information leaflet outlining the study and what participation would involve. Following full written informed consent, recruitment took place from 14^th^ December 2023 until 30^th^ June 2024. Data collection was based on previous work to include a purposively recruited large, maximum variety sample which offered adequate credibility, rather than recruiting to data saturation.

In each activity, we separated time in which the questions were focused on either CHIM in PLHIV or CHIM using mycobacteria (BCG or TB CHIM). We allowed for a rest break of at least 30 minutes in-between topic discussions. We collected data using topic guides. We iteratively amended the guides during the study to explore emerging topics. Each FGD lasted approximately 2 hours, and each IDI about one hour. Field notes were collected and kept in a “reflexive diary”. We audio-recorded both FGDs and IDIs, uploaded them on a secure server in REDCap, then transcribed the recordings verbatim. Discussions conducted in Chichewa were transcribed, then translated into English. Interviews were conducted by a female research assistant with experience in qualitative research who had no previous experience of the interviewed study participants.

### Data analysis

Transcripts were organised using NVivo 14 and thematically analysed using thematic and framework analysis methodology with broadly deduced themes (recruitment, practical or study-specific considerations) and sub-themes were inductively derived (CG, ACG)[20]. Consensus was reached on themes by AG and CG during regular analysis meetings, and later by all members of the team.

## Results

### Participants and recruitment discussions

A total of 115 participants were contacted and 114 participated, all with written informed consent. Demographic details are summarised in Table 1. Fourteen FGDs were carried out with a purposively recruited, maximum variety sample of 106 participants[21]; PLHIV recruited from clinics (9), former TB patients recruited from the TB registry (7), healthy adult community members identified by recruitment of volunteers through Community Advisory Groups (CAGs) (19), former participants of previous CHIM studies approached by phone or text message (14), community opinion leaders (local chiefs) (5), religious leaders (6), and CAG members (13) recruited through our ongoing public engagement, HIV care clinicians and nurses (10), TB care clinicians and nurses (13), and Health Surveillance Assistants (HSAs) specialising in HIV or TB care (10) recruited through local health services. We conducted separate FGDs for each of these groups to account for possible culturally oriented power dynamics, but we combined men and women in the groups. We also carried out eight IDIs focused on safety and ethical considerations, with HIV clinician scientists (2), tuberculosis clinician scientists (2) and local Research Ethics Committee members (4).

**Table 1.** Participants demographics.

| Characteristic | CATEGORY |  |  |  |  |  |  |  |  |  |  |  |  |
| --- | --- | --- | --- | --- | --- | --- | --- | --- | --- | --- | --- | --- | --- |
|  | Overall<br>N = 114<sup>1</sup> | CAG<br>N = 13<sup>1</sup> | Community<br>leader<br>N = 5<sup>1</sup> | Community<br>member<br>N = 19<sup>1</sup> | Former<br>CHIM<br>N = 14<sup>1</sup> | Former<br>TB<br>patient<br>N = 7<sup>1</sup> | HIV Care<br>Provider<br>N = 10<sup>1</sup> | HSA<br>N = 10<sup>1</sup> | PLHIV<br>N = 9<sup>1</sup> | Policy<br>maker<br>NCST<br>N = 2<sup>1</sup> | Religious<br>Leader<br>N = 6<sup>1</sup> | Senior<br>Doctor<br>N = 6<sup>1</sup> | TB Care<br>provider<br>N = 13<sup>1</sup> |
| GENDER |  |  |  |  |  |  |  |  |  |  |  |  |  |
| Female | 65 | 7 | 0 | 13 | 7 | 5 | 3 | 8 | 7 | 1 | 3 | 3 | 8 |
| Male | 49 | 6 | 5 | 6 | 7 | 2 | 7 | 2 | 2 | 1 | 3 | 3 | 5 |
| AGE | 43 (35, 49) | 48 (43, 52) | 43 (43, 49) | 46 (38, 67) | 29 (28, 33) | 47 (45, 51) | 31 (30, 40) | 47 (41, 51) | 47 (45, 50) | 44 (35, 53) | 54 (43, 57) | 46 (36, 48) | 38 (31, 39) |
| INTERVIEW_TYPE |  |  |  |  |  |  |  |  |  |  |  |  |  |
| FGD | 106 | 13 | 5 | 19 | 14 | 7 | 10 | 10 | 9 | 0 | 6 | 0 | 13 |
| IDI | 8 | 0 | 0 | 0 | 0 | 0 | 0 | 0 | 0 | 2 | 0 | 6 | 0 |

<sup>1</sup> n; Median (Q1, Q3)

### Thematic analysis

Thematic analysis identified four primary themes, and their sub themes as illustrated in Table 2. The primary themes were: (1) General views of CHIMs, (2) CHIM recruitment and consent, (3) CHIM study practical considerations including gender issues (4) Individual CHIM study specific considerations. Each theme, and the evidence to support it, are presented in order.

**Table 2.** Key Themes and sub-themes.

| Theme | Sub-themes |
| --- | --- |
| Acceptability of advanced CHIMs | Knowledge of research or participation in previous research<br>Recognised clinical need<br>National or community interest |
| CHIM recruitment factors | Misinformation<br>Community engagement strategies<br>Informed consent and literacy<br>Gender-specific factors and contraception<br>Poverty and financial compensation |
| CHIM study implementation factors | Access to health care<br>Residential stay – gender roles and community perception |
| Study specific factors for PLHIV | Vulnerability<br>CD4 count |
| Study specific factors for TB CHIM | Biopsy and use of tissue<br>Bronchoscopy<br>Familiarity with BCG |

#### General views of CHIMs

Participants across the focus group discussions and in-depth interviews understood the scientific potential of CHIM to contribute to clinical research work and development of new drugs and vaccines. Participants were supportive of proposals to conduct CHIM studies in Malawi. Many participants felt the anticipated benefit outweighed the potential risks (Figure 1a).

**Figure 1.**
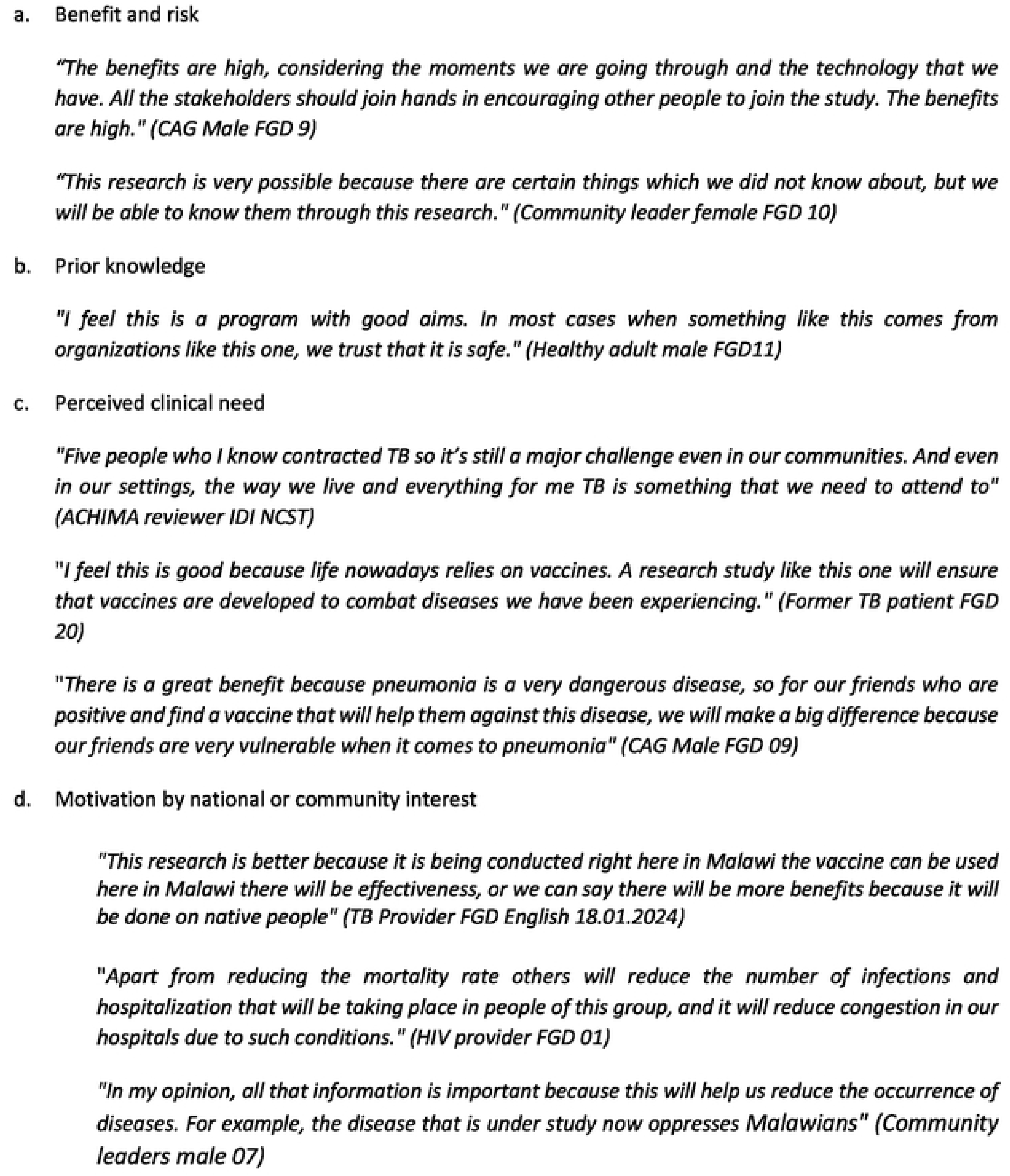
Primary Theme 1 General acceptability of CHIMs

Participants with prior knowledge or participation in previous research expressed specific confidence in research conducted by the Malawi-Liverpool Wellcome Program, drawing from their prior experience with previous studies including community vaccine studies (community members), surveys and CHIM studies (mostly university students). Prior involvement appeared to build trust in both the research process and outcomes (Figure 1b).

Participants perceived an acute need for improving treatment and prevention of pneumococcal disease among people living with HIV. Tuberculosis (TB) was seen as a priority disease for intervention in Malawi and this influenced participants from both focus group and in-depth interviews. Their reflections were focused on personal loss as well as broader national burden. Many felt that CHIM studies were justified due to this clinical need, and research was viewed as one potential solution to health challenges Malawi is facing (Figure 1c).

Participants were motivated by national or community interest and viewed participation in CHIM studies as a personal sacrifice, recognising the benefits of the research to the wider community. Participants believed that CHIM research could strengthen the local health sector and benefit future patients. A strong sense of national pride and belonging underpinned a belief that locally conducted research could foster trust in medication and vaccine development (Figure 1d).

#### Perspectives on CHIM recruitment

Most participants supported the development of context-specific research volunteer recruitment strategies including briefing community leaders, posting on social media, public events and advertising on local radio and TV. They articulated that this approach would address cultural and site-specific challenges and ensure that participants’ recruitment was both ethical and safe. The sub-themes discussed were community engagement, literacy, misinformation, gender specific factors, poverty, compensation and incentives to research participation.

Participants highlighted the importance of mass awareness campaigns stating that effective communication efforts could contribute significantly to the likelihood of CHIM studies being well received and accepted by community members. Stakeholders recommended use of mass communication channels such as megaphones and radio adverts as effective tools for broad community outreach alongside partnerships between trusted community figures and research outreach workers (Figure 2a).

**Figure 2.**
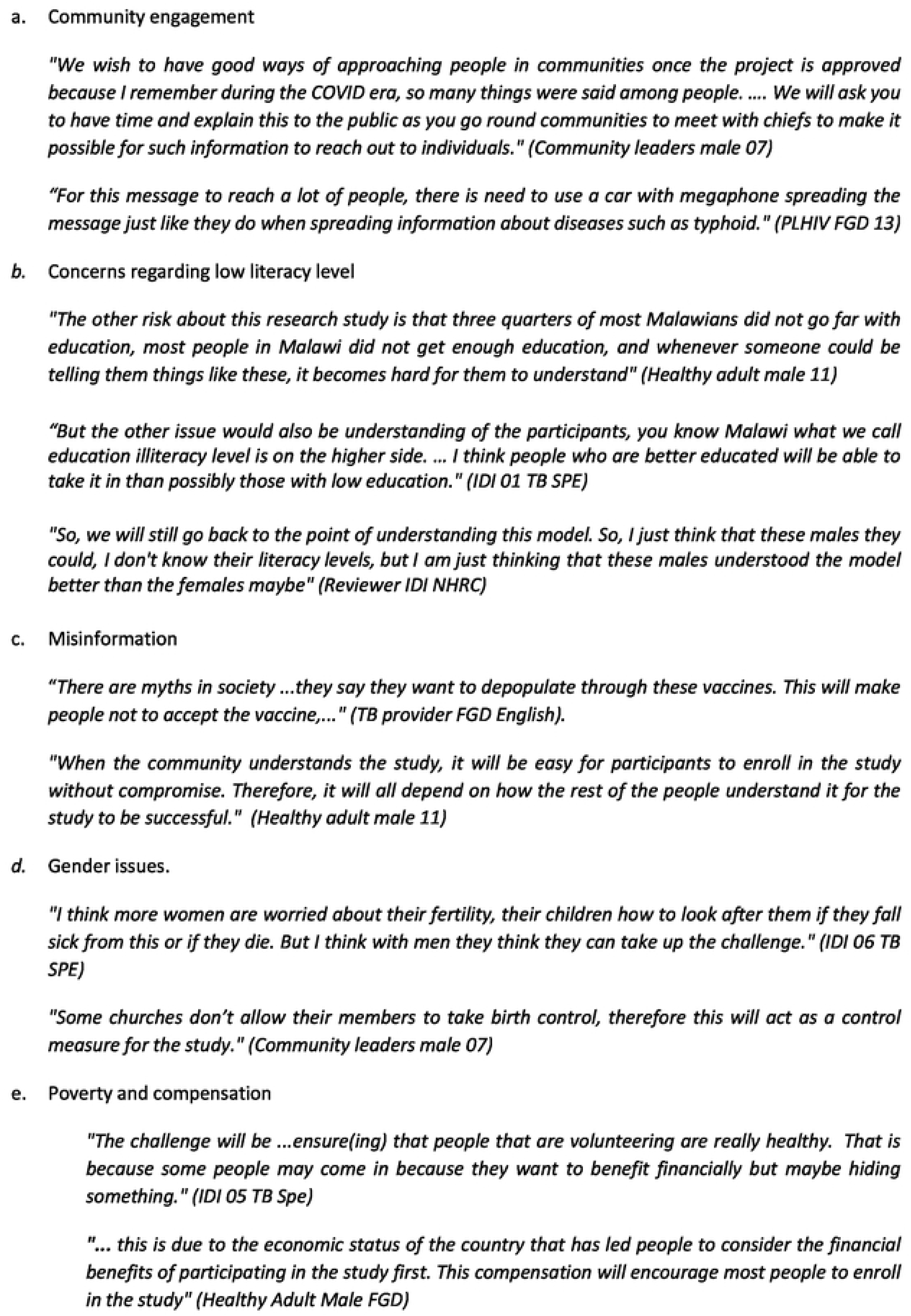
Evidence in Primary Theme 2: Malawi-specific CHIM study volunteer recruitment factors

Concerns were raised by participants regarding low literacy levels among the community, and the complexity of CHIM studies which may pose challenges in obtaining fully informed consent. Both FGD and IDI participants noted that recruitment might be more successful among individuals with at least a secondary education, and potentially favour men, as they were perceived to have higher literacy levels. Participants in focus group discussions, however, expressed a good understanding of CHIM in PLHIV, or CHIM using mycobacteria, and agreed that while low literacy might be a challenge, this did not constitute a complete barrier to participation (Figure 2b).

Misinformation was widely discussed as a significant barrier to recruitment and participants discussed experiences in the COVID pandemic and subsequent vaccine roll out, including rumours that the vaccination was a population control measure being inflicted upon Malawian people. Some rumours originated from social media (rumours of control) while others were more local (vaccination can lead to cholera). Community members understood the impacts of persistent misinformation on both direct study recruitment and the risks of community misunderstanding (Figure 2c).

Participants anticipated that recruitment would be more challenging for women than men. Key concerns were the potential impact of study participation on childcare responsibilities, and the study requirement for contraception during study period (agreement to use contraception is an inclusion criterion in CHIM studies). Participants agreed that women enrolled in CHIM studies should have a contraceptive plan in place for the study duration viewing this as a necessary safety measure. Male contraception was not discussed as the groups did not perceive a specific risk to males or their unborn offspring associated with procreation during CHIM participation (Figure 2d).

Participants highlighted the complexity of determining appropriate compensation in a resource-poor context. Participants stated that compensation decisions should be influenced by perceived risk factors, family well-being and cultural expectations. Participants recognised that Malawi’s ongoing economic hardships further complicate the assessment of what constitutes fair and ethical compensation (Figure 2e).

#### Practical considerations for implementing CHIM studies

Adapting CHIM studies to ensure alignment with the socio-economic and cultural context in Malawi emerged as a key consideration in IDIs and in FGDs. Participants raised significant concerns regarding limited access to medical care in Malawi and how that would influence CHIM study implementation. They also discussed residential participation in the study, gender issues related to residential stay, and community perceptions of residential stay.

Participants emphasized the importance of ensuring that individuals taking part in a CHIM study could access quality health care both during and after the study period. They emphasized the need for clear and comprehensive health care provision as part of study design to ensure participants’ safety and ethical standards. Participants recommended that health care support should not be limited to study period but should safeguard long-term well-being of participants (Figure 3a).

**Figure 3.**
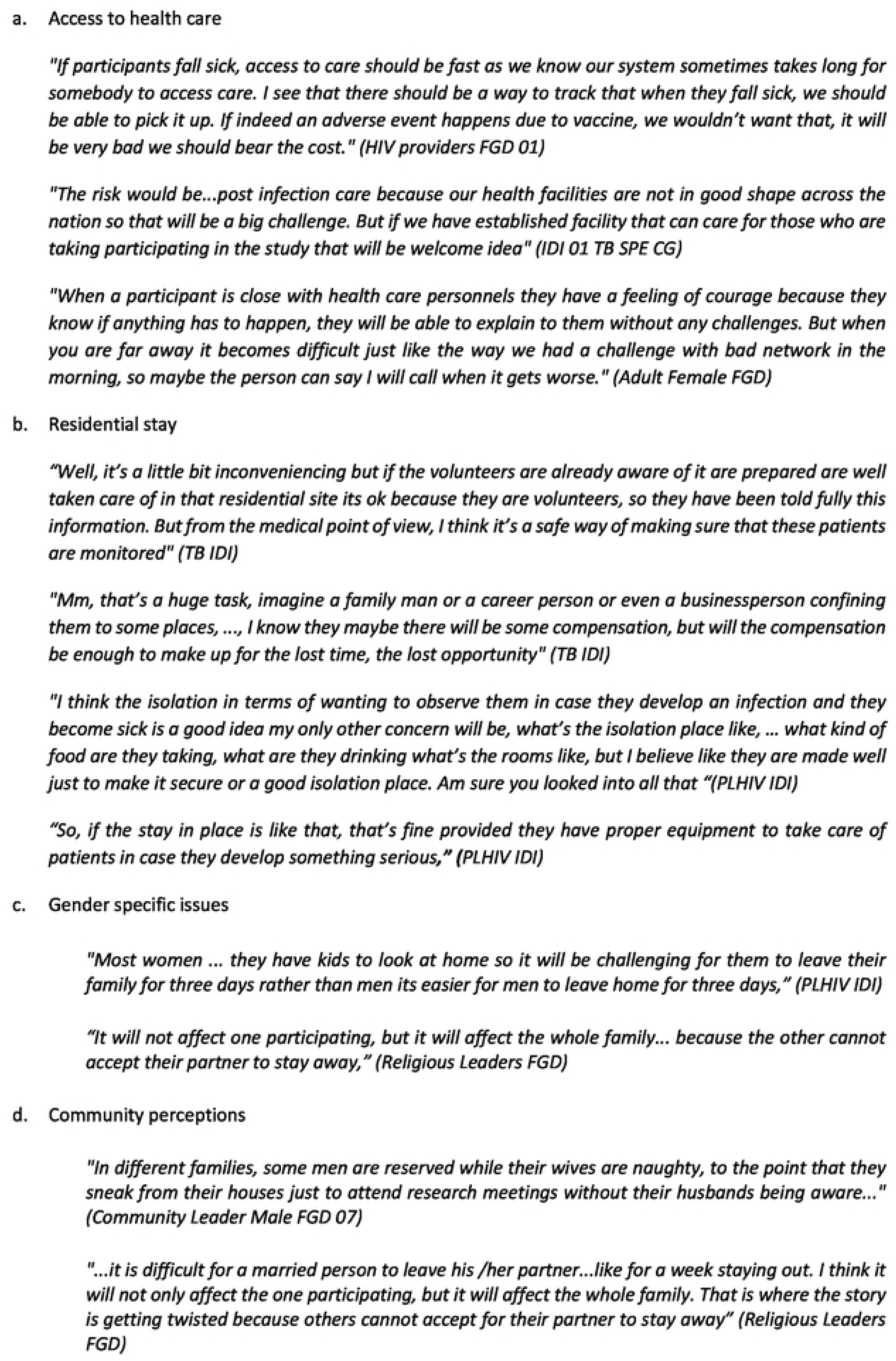
Primary Theme 3: Malawi- specific CHIM study implementation factors

Residential stay in a conference centre to ensure easy access to medical support, but not constituting inpatient medical supervision, was supported by most participants. This could also be viewed as an inconvenience, however, leading to loss of income during the stay. To address the concerns raised participants emphasized the need for the study site to be adequately equipped and include provision of essential needs including food and water to support participants’ well-being during the stay (Figure 3b).

Gender specific issues discussions were in relation to residential stay in the context of childcare roles and family life acting as a barrier to participation for female participants particularly married ones. This was not felt for male participants (Figure 3c).

Participants expressed concern that community perceptions might result in reputational damage to individuals, especially women, who would stay away from home for study activities. They noted it would be difficult for participants to enroll in the study without the wider community being aware of leading to stigma or gossip. Participants emphasized the importance of strong and transparent communication to build understanding and promote acceptance of study (Figure 3d).

#### Perspectives of participants on Pneumococcal CHIM in PLHIV and TB CHIM

Two specific conversations were conducted with each IDI and FGD. The first topic was discussion of a CHIM to develop pneumococcal carriage in PLHIV. The second topic was a CHIM to study immunological responses to either skin or lung BCG instillation, intended in HIV negative healthy adults. Participant concerns were separately discussed in the context of conducting a Pneumococcal CHIM in PLHIV or a TB CHIM in healthy adults.

Regarding the pneumococcal CHIM in PLHIV, participants expressed concern about the clinical vulnerability of people living with HIV, recognizing them as being at high risk of infections. They emphasized the need for caution when considering the inclusion of this group in CHIM studies. Suggestions were made to ensure that the study protocol adequately addresses these risks, including thorough health assessments based on CD4 count to ensure that only healthy individuals were enrolled to minimize risks (Figure 4a).

**Figure 4.**
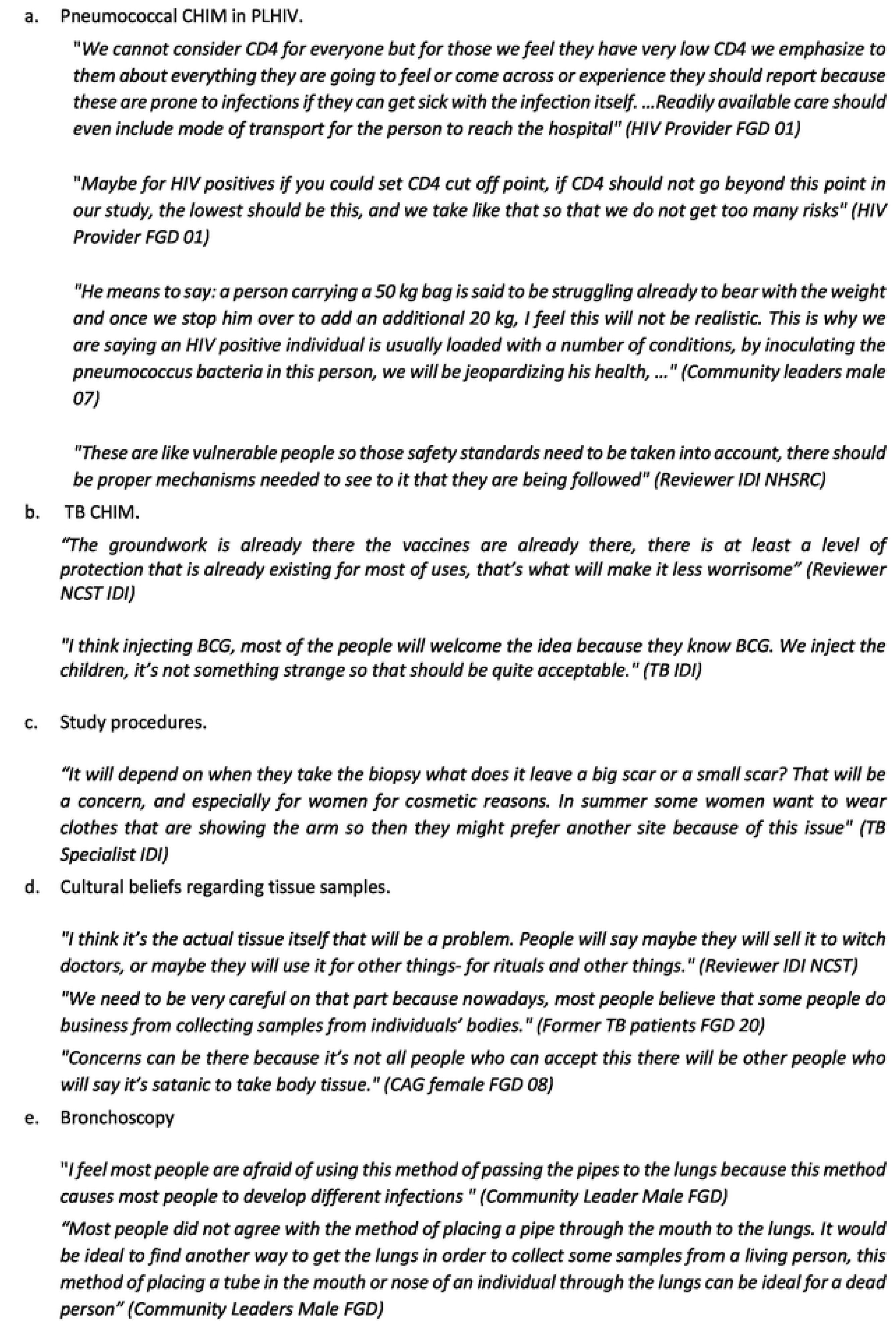
Primary Theme 4: Individual CHIM study specific considerations

In contrast, there was broad support for the TB CHIM from stakeholders, largely due to familiarity with the BCG vaccine which was proposes as the infectious agent under study. BCG is well known as part of the routine childhood immunization programme in Malawi. Additionally, stakeholders recognized the significant burden TB places on the Malawian population and expressed hope that such studies could contribute to reducing TB cases in the long term (Figure 4b).

Participants expressed specific reservations about certain study procedures in the proposed BCG CHIM. The skin biopsy procedure to determine infectious load raised concerns related to cosmetic outcomes—especially among women. Participants associated the biopsy with potential pain, discomfort, and the possibility of scarring. Additionally, there were worries about delayed healing and the risk of infection at the biopsy site, which contributed to hesitancy about this aspect of the study (Figure 4c).

In addition to physical concerns there were notable fears rooted in cultural beliefs and misinformation. Some participants associated the collection of human tissue with local myths or suspicions about its possible use in rituals or harmful practices. These misconceptions contributed to anxiety and mistrust around biopsy procedures. Participants emphasized the importance of transparent communication about the purpose of tissue collection, and how samples would be handled after use. Clear messaging on these points was seen as critical to optimise community trust and acceptance of study procedures (Figure 4d).

Bronchoscopy and bronchoalveolar lavage were discussed as a means of evaluating lung immunity. This was viewed by participants as being particularly invasive as a research procedure in the absence of a clinical indication. Many expressed concerns about the potential risk associated with the procedure, including infection, discomfort, and the possibility of complications (Figure 4e).

## Discussion

This study distilled topics and opinions found among a large, diverse group of Malawian stakeholders regarding CHIM studies at a time when studies either recruiting at-risk participants or trialling more complex pathogens are being developed. There were both expected and unexpected findings in each of the four primary themes that emerged in thematic analysis.

A strength of this study is its inclusivity of a broad range of study participants from different backgrounds increasing its transferability. Additionally, findings were subject to rigorous interpretive challenge during analysis. The study was designed by Malawian researchers with experience in human challenge experiments, interviews were conducted by a research assistant (CG) not directly involved in the conduct of CHIM, analysis was led by an “outsider” not involved in any of our work (AG), the combination of which allowed for greater reflexivity. Limitations included inherent weakness of focus group discussions as they lack individuality of voices, and in-depth interviews were exclusively with experts.

CHIM studies have been conducted in Malawi for 7 years, so a level of acceptance was expected, particularly among those with direct experience of the programme[22, 23]. An unexpected finding was the trust placed in the MLW programme, and this mirrors data from Kenya where the KEMRI/Oxford brand is very strong[24]. The positive opinion of the research programme contrasted starkly with opinion regarding local government health services. This resonated with the authors experience and current reporting in the Malawi press about public health services. Discussion of the motivation of volunteers resonates with experience from other parts of the world, including the UK during the COVID pandemic when the SARS-CoV CHIM was developed[25].

The importance of public awareness expressed aligned with our previous work showing that recruitment is mainly from word of mouth[26], possibly supported by community information programmes. We use radio phone-in programmes, live TV debate and public meetings to disseminate information, but most volunteers present after contact with friends who are previous volunteers.

Comprehension is important to facilitate informed consenting in CHIM studies. Our local ethics review board requires that recruitment should be conducted in either English (often the preferred language of those with secondary education) or Chichewa, the national language. Limited community understanding and misinformation led to low COVID vaccine uptake in Malawi (11%) but established infant vaccination programmes are trusted and have more than 90% coverage. This highlights the value of this study to understand nuanced participants perceptions and improve understanding with informed voluntary participation in CHIM studies.

FGD participants expected that women might be less likely to recruit than men. Our lived experience is that the opposite is true which may be due to the age demographic (university students) of many of our participants making it unusual for us to recruit women with caring responsibilities. In our published CHIM studies testing pneumococcal vaccines, more women than men were recruited in Malawi, but more men in Liverpool, UK. Our early feasibility work recruiting PLHIV from anti-retroviral therapy (ART) clinics found that 75% of recruited volunteers were women. We attribute some of this to the reluctance of men to attend clinics but even using other recruitment settings, it has proved challenging to recruit men[27]. This gender imbalance highlight the need to further explore context-specific recruitment strategies targeting men to ensure inclusive participation in CHIM studies.

Appropriate compensation for research participation is very important. We have published specifically on this topic and use our own local guideline when estimating compensation for out-of-pocket expenses, travel, loss of earnings and discomfort[28]. There is no perfect solution to this challenge, but frequent review of compensation offered, and rigorous screening to avoid over-recruitment are important. The introduction of biometrics for identification is helpful in preventing over-volunteering. Thorough community engagement, transparent communication and culturally sensitive implementation are also essential to improve community understanding of biometrics as a protective measure rather than a threat.

One motivation for research volunteering in Malawi has been shown to be access to health care[29]. This was re-emphasised in this study, now more than 20 years after our first publication. Despite the relative abundance of doctors graduating from local medical schools, health care provision has not kept pace with population growth and access to health care remains a pressing problem in Malawi. It is therefore essential to address such ‘therapeutic misconceptions’ and ensure voluntary informed participation in CHIM studies to promote public health.

Residential care for the period immediately following experimental inoculation is reassuring for participants, staff team and regulators in our context. Some concerns were raised in focus groups regarding community perceptions about this residential approach, particularly regarding reputational damage for women. Our experience, however, has been that the feedback from participants in pneumococcal challenge studies, and early CHIM studies recruiting PLHIV, has been universally positive. We have very recently[30] started piloting BCG CHIM which is conducted as a soley outpatient study.

Community perceptions on PLHIV participation in CHIM in a context where 11% of adults are taking ART were critical to this study. Clearly the message regarding good viral suppression and healthy living with HIV have not turned the community dialogue away from the immediate association of HIV infection with AIDS that was so prevalent 20 years ago[30]. It is of course appropriate that our current and planned studies align with the expectations of the focus group participants that CD4 count/viral suppression are necessary pre-requisites of entry into these studies.

The participants enthusiasm for TB research was a clear finding in this study. TB is a feared disease in Malawi, and the failure of BCG to protect adults is widely understood. It is therefore important to recognise this community priority and conduct research on TB vaccines that may protect adults. The implementation of BCG-CHIM studies is therefore essential to accelerate the development of TB vaccines that not only safeguard adult populations but also help mitigate the emergence of drug-resistant TB over time.

Specific concerns about biopsy, tissue collection and bronchoscopy were expected, and it is reassuring to see that the pragmatic concern about scarring, as well as reporting of community superstition about tissue misappropriation came up in discussions. The concern about bronchoscopy was surprising as our programme has a 28-year history of research bronchoscopy spanning many projects and feedback has been positive, including many repeat volunteers[29]. This finding indicates that some additional community discussion specifically about bronchoscopy would be of value in the community engagement programme.

In summary, this extensive consultation exercise has confirmed community support for locally relevant research, along with raising appropriate concerns about recruitment, safety, consent, information, and remuneration in CHIM. The participants were able to confirm a respect for volunteering in research based on community benefit, even when the studies suggested involved both subjects with increased susceptibility to infection, and pathogens of special interest.

## Data Availability

Full dataset is available on application to the Head of the Data unit at MLW. Alfred Muyaya

